## Supplementary Information for "Association between mobility, non-pharmaceutical interventions, and COVID-19 transmission in Ghana: a modelling study using mobile phone data"

**Supplemental Section 1**

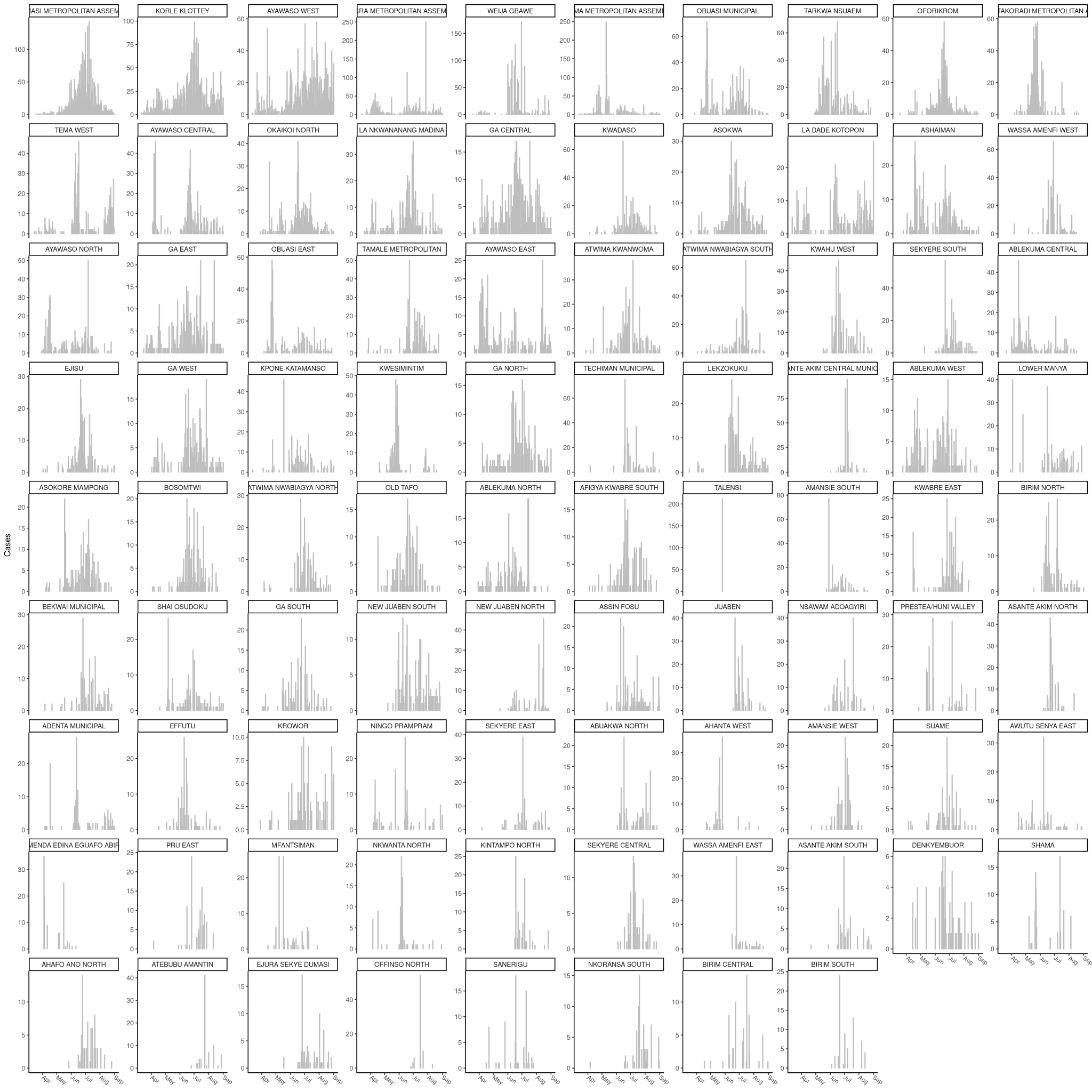

**Supplemental Figure 1. *Reported COVID-19 cases in individual districts.*** *The daily number of reported COVID-19 cases through time, aggregated from line list data to individual districts.*

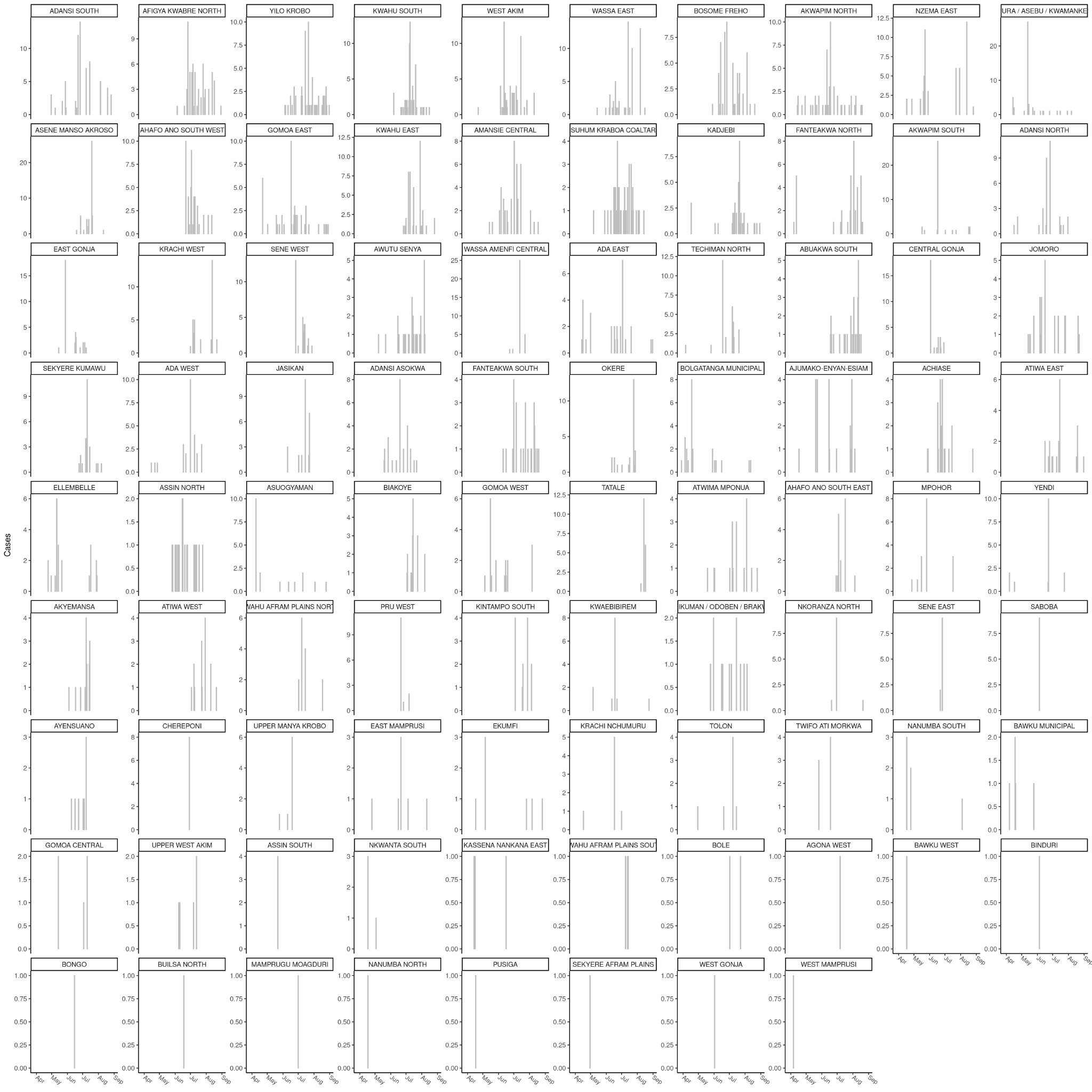

***Supplemental Figure 1 (continued). Reported COVID-19 cases in individual districts.*** *The daily number of reported COVID-19 cases through time, aggregated from line list data to individual districts.*

| **District** | **Date** | **Removed** |
| --- | --- | --- |
| Ayawaso West | 2020-08-01 | 1115 |
| Ayawaso West | 2020-07-30 | 334 |
| La Nkwananang Madina | 2020-08-06 | 426 |

***Supplemental Table 1. Manual reporting outliers.*** *Outliers replaced by linear interpolation between the preceding and following records following visual inspection of aggregated case counts in individual districts. Original reported cases for these dates exceeded 5 times the number of cases reported on all previous dates.*

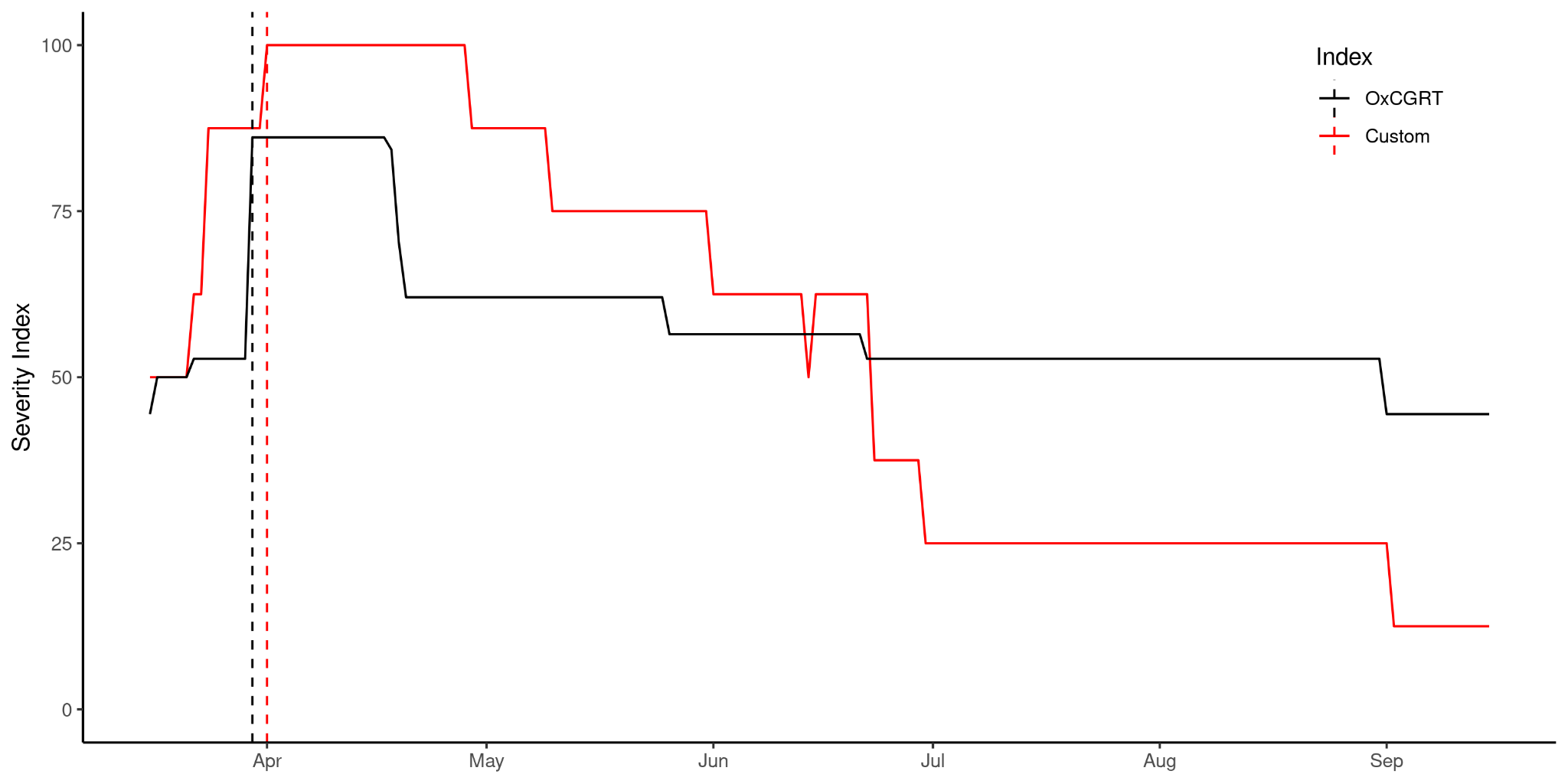

***Supplemental Figure 2: Comparison of Stringency Indices.*** *A comparison of the OxCGRT and custom stringency indices. Dashed lines indicate the dates of maximum NPI stringency. Both indices reflect similar patterns in the progression of NPI stringency in Ghana and identify the similar dates of maximal NPI stringency.*

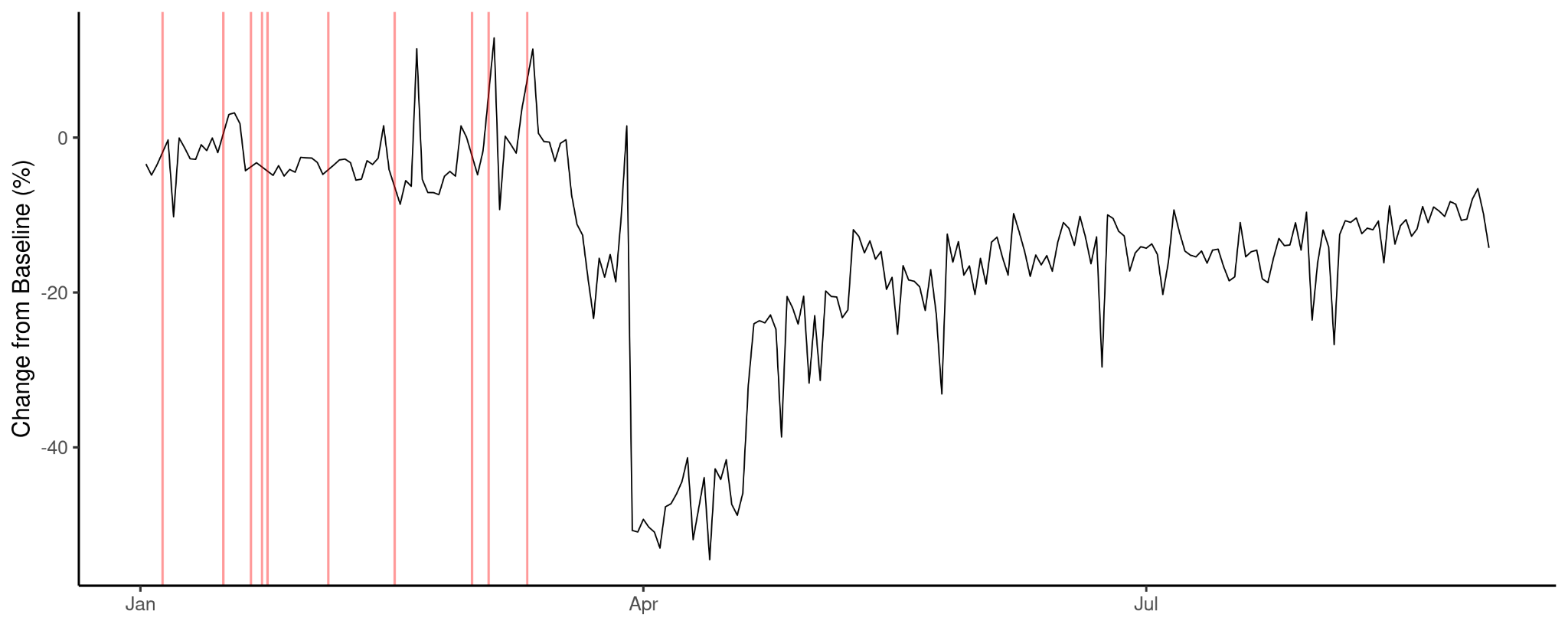

***Supplemental Figure 3. Missing dates in Vodafone mobility data.*** *The missing dates in Vodafone Mobility data (red lines) and Vodafone mobility data in Accra Metropolitan Area. Missing dates in Vodafone data were the same for all districts. Missing dates comprised 4.1% of the entire time series. Linear interpolation was used to impute data for missing dates.*

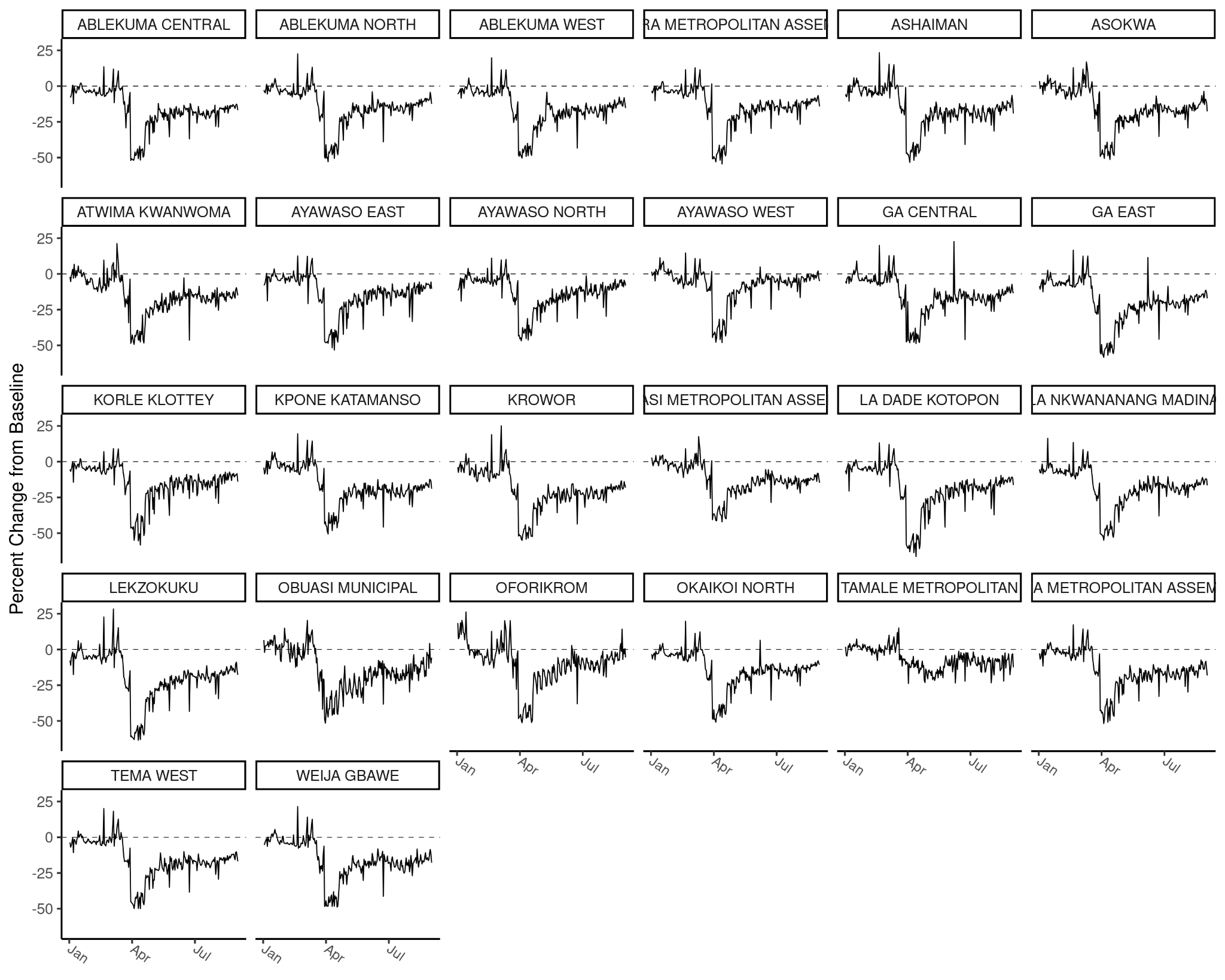

***Supplemental Figure 4. Vodafone mobility indicator in different districts.*** *The mobility indicator collected by Vodafone Ghana in individual districts. Measured as a percent change from baseline values.*

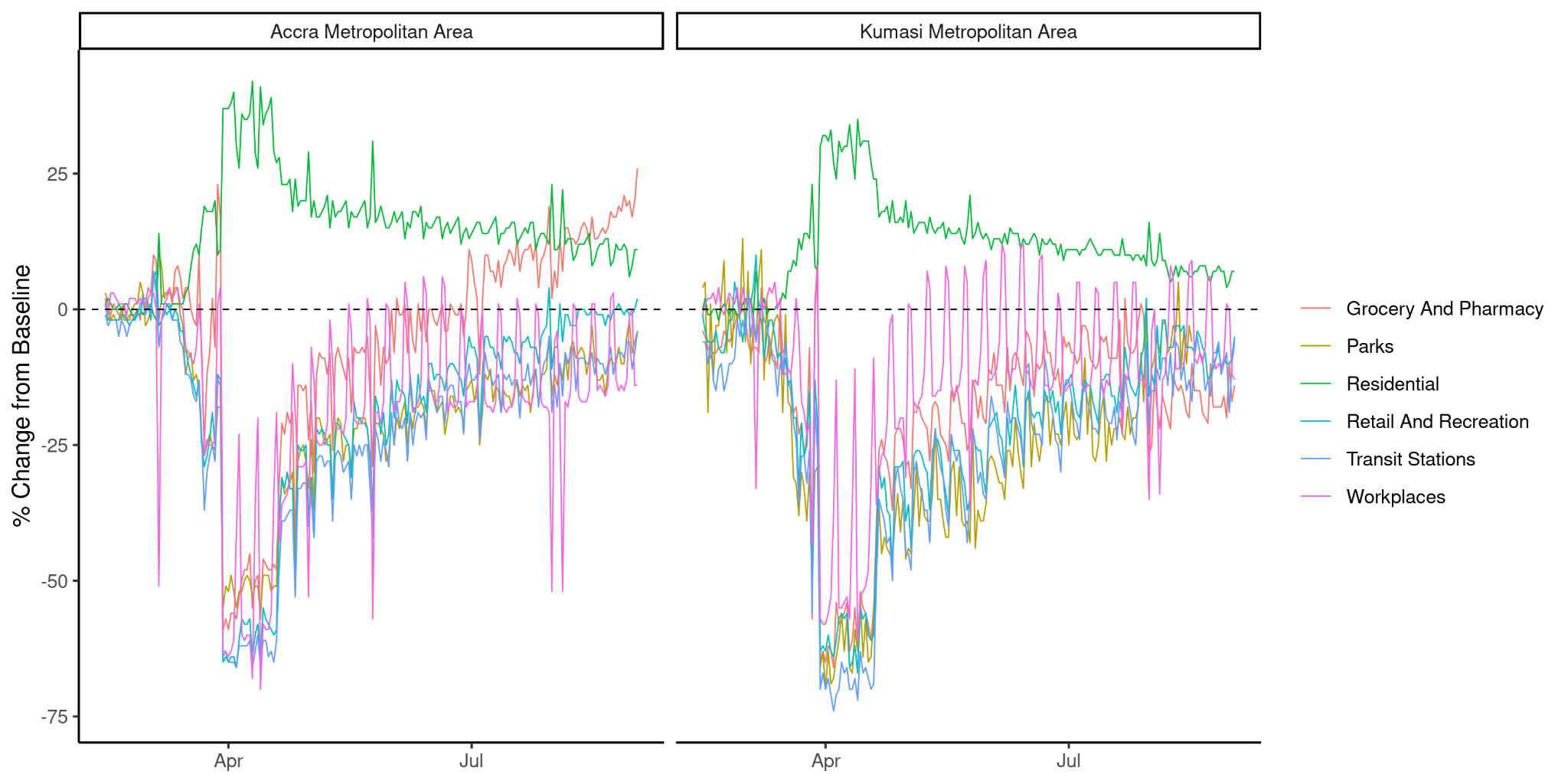

***Supplemental Figure 5. Google mobility indicators in different settings****. Google mobility indicators in all settings in Accra and Kumasi Metropolitan areas.*

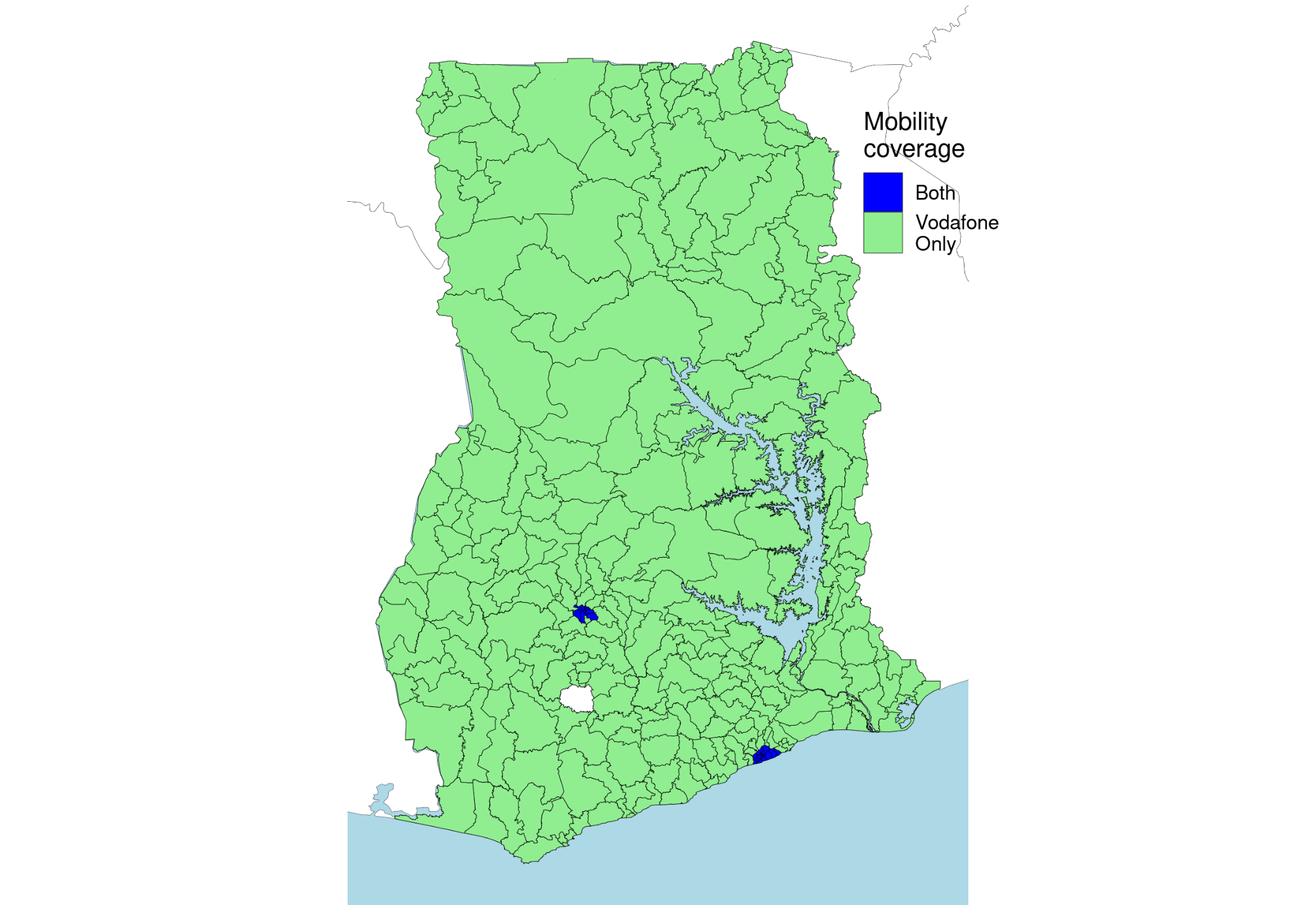

***Supplemental Figure 6. Availability of mobility indicators.*** *The availability of mobility data from Vodafone Ghana (“Vodafone Only”) and Google (“Both”). Google mobility data is only available for Kumasi and Accra metropolitan areas.*

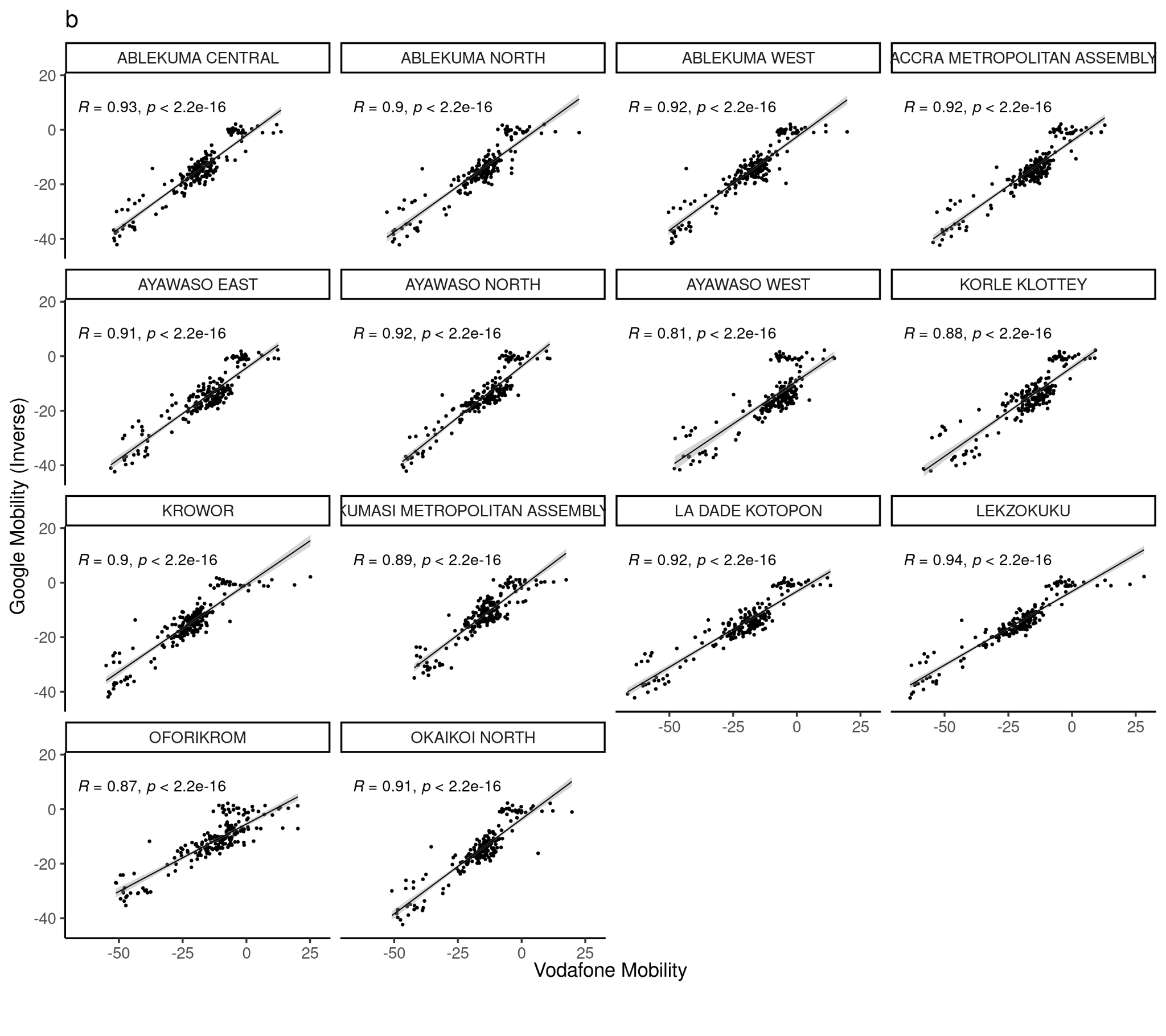

***Supplemental Figure 7. Association between Google and Vodafone mobility indicators.*** *The association between Google and Vodafone mobility indicators in the Accra and Kumasi metropolitan areas between 12th March and 1st September 2020. Note that these data are collected from two different sources (Google: GPS, Vodafone: CDRs) and describe different aspects of mobility (Google: activity in “residential” areas, Vodafone: travel between administrative districts).*

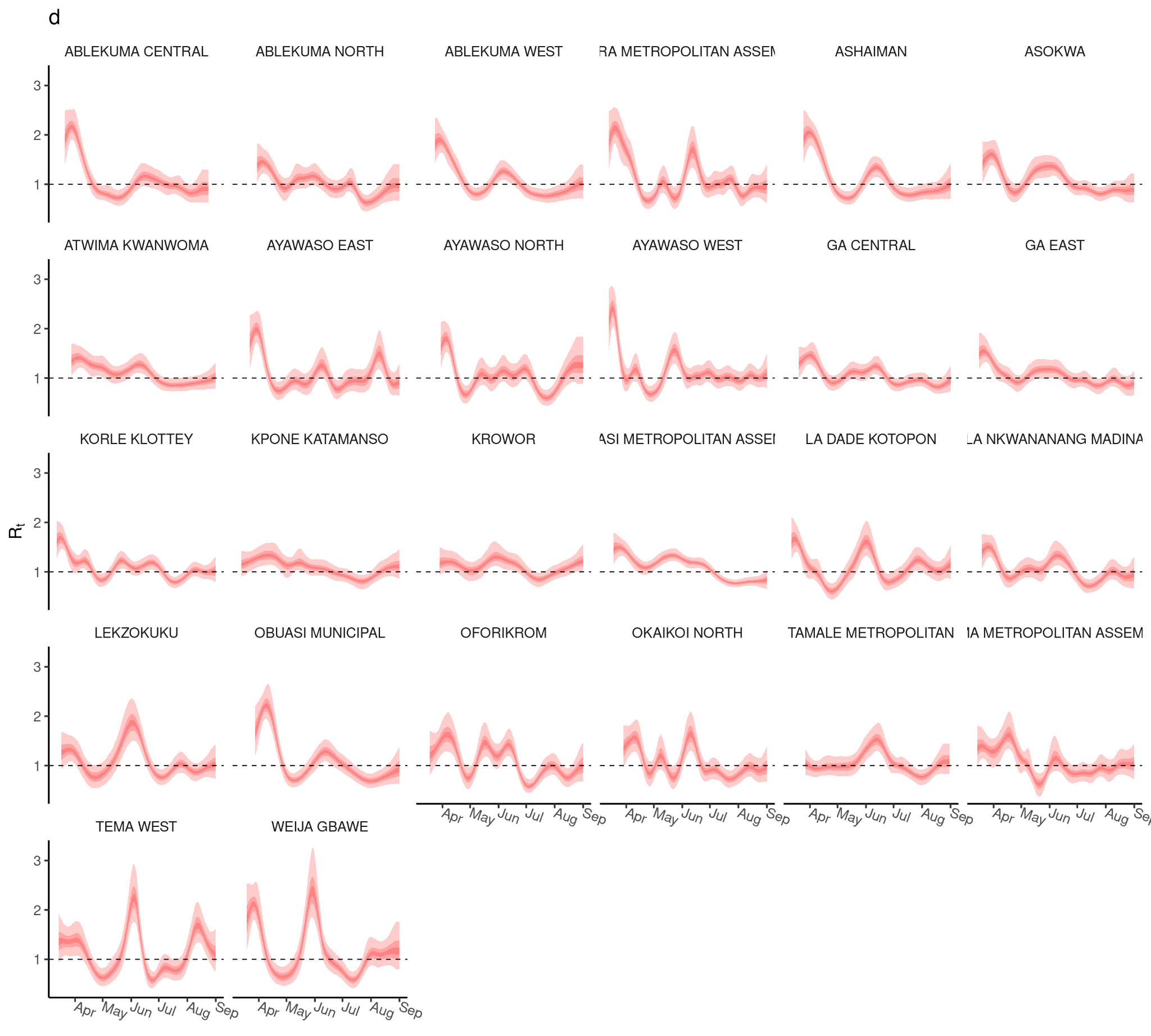

***Supplemental Figure 8. R_t_ estimates for individual districts.*** *Estimates of R_t_ in individual districts.*

| **Predictors** | **Estimates** | **CI** | **p** |
| --- | --- | --- | --- |
| (Intercept) | -0.000 | -0.084 – 0.084 | 1.000 |
| NPI | 0.769 | 0.754 – 0.783 | <0.001 |

***Supplemental Table 2. Results of the Level 1 model.*** *The results of the level 1 model of Vodafone Mobility and the (inverse) custom NPI indicator. This shows a positive association between mobility and inverse NPI stringency. The residuals of this model were used in the Level 2 model.*

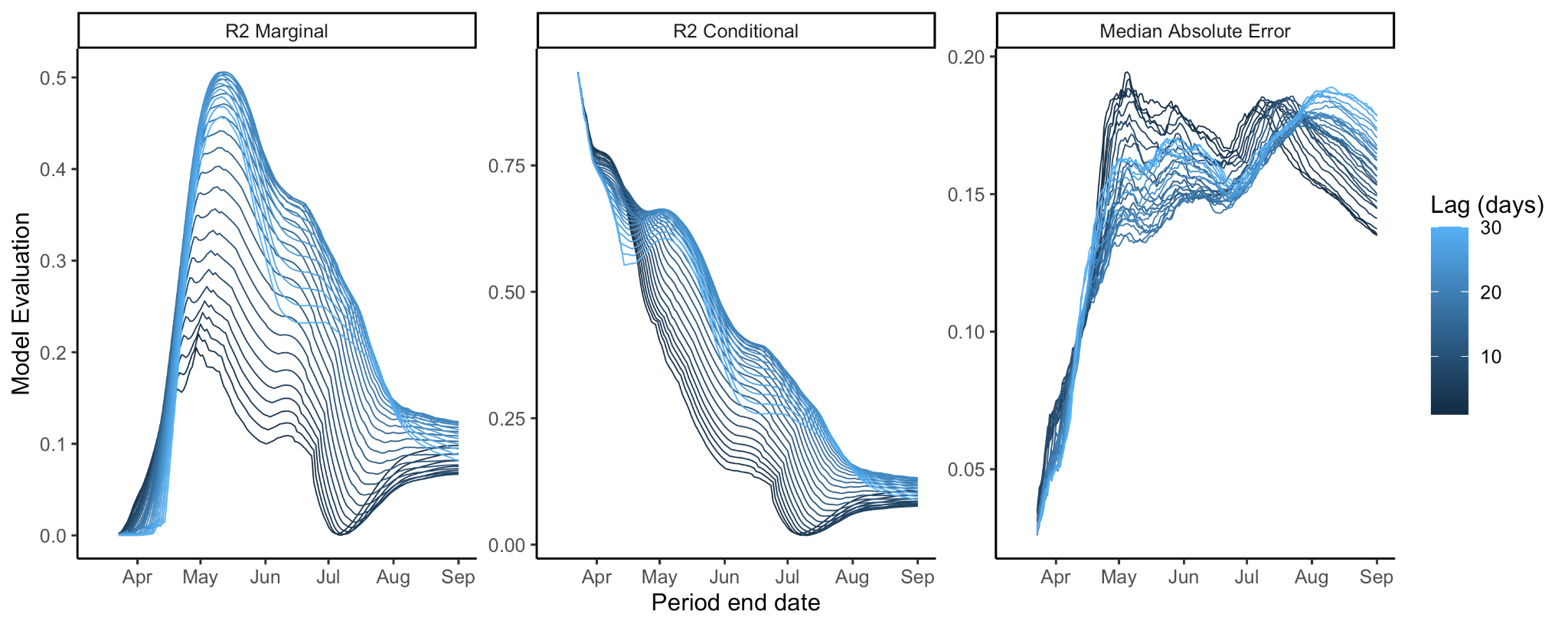

***Supplemental Figure 9. Model performance for different periods.*** *The performance of the multilevel model through time, measured by marginal and conditional R^2^ (trained on periods beginning March 12th and ending from March 19th to September 1st).*

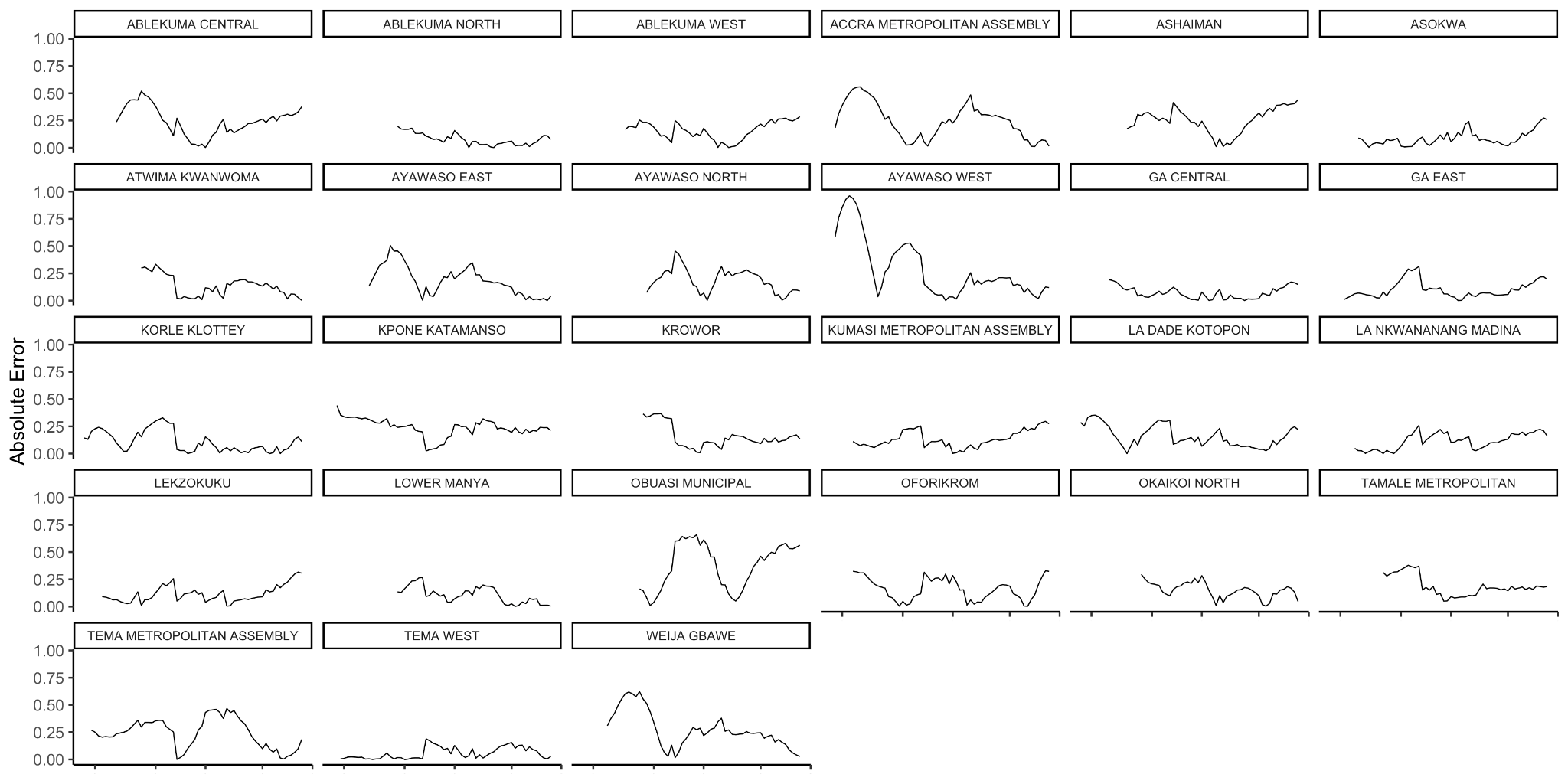

***Supplemental Figure 10. Absolute error by date and district.*** *The absolute error of model estimations on each date per district. For some districts, we observe high model error after the first reported cases.*

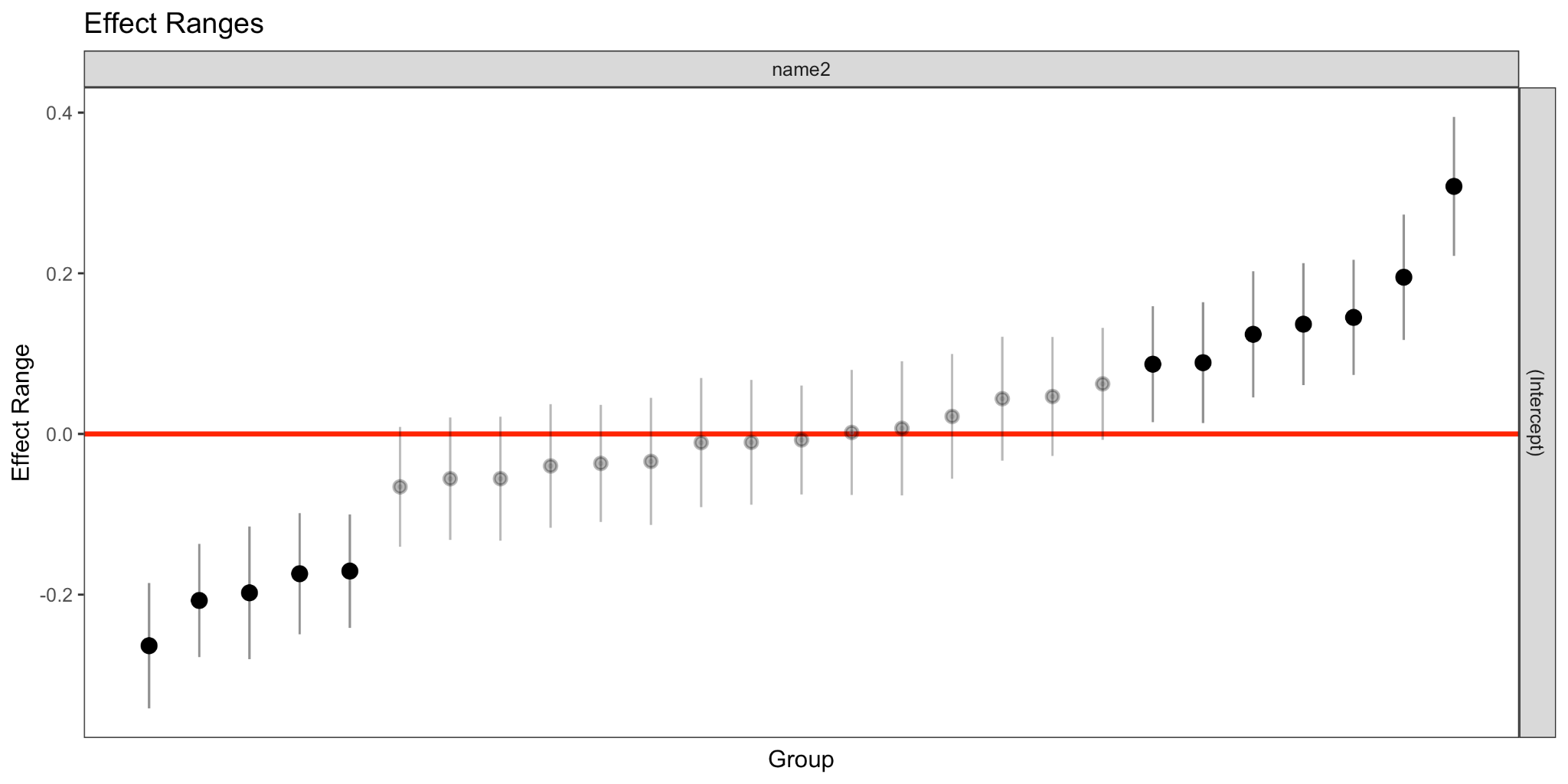

***Supplemental Figure 11. Random effect sizes for a model with truncated time series.*** *The district-specific random effect sizes for the model trained between 12th March and 15th May. Ten random effects are distinguishable from 0.*

**Supplemental Section 2**

Sensitivity analysis of model outputs using the Vodafone Mobility Indicator and OxCGRT stringency index. This analysis shows similar results to the model using Vodafone data and the custom stringency index with an increase in the model’s explanation of R_t_ variance early in the epidemic and subsequent decrease after June 2020 (Supplemental Figure 11). Inclusion of the OxCGRT NPI index explains a greater amount of variance in R_t_ later in the epidemic. In this analysis we identified an optimal lag of 22 days (Supplemental Figure 12).

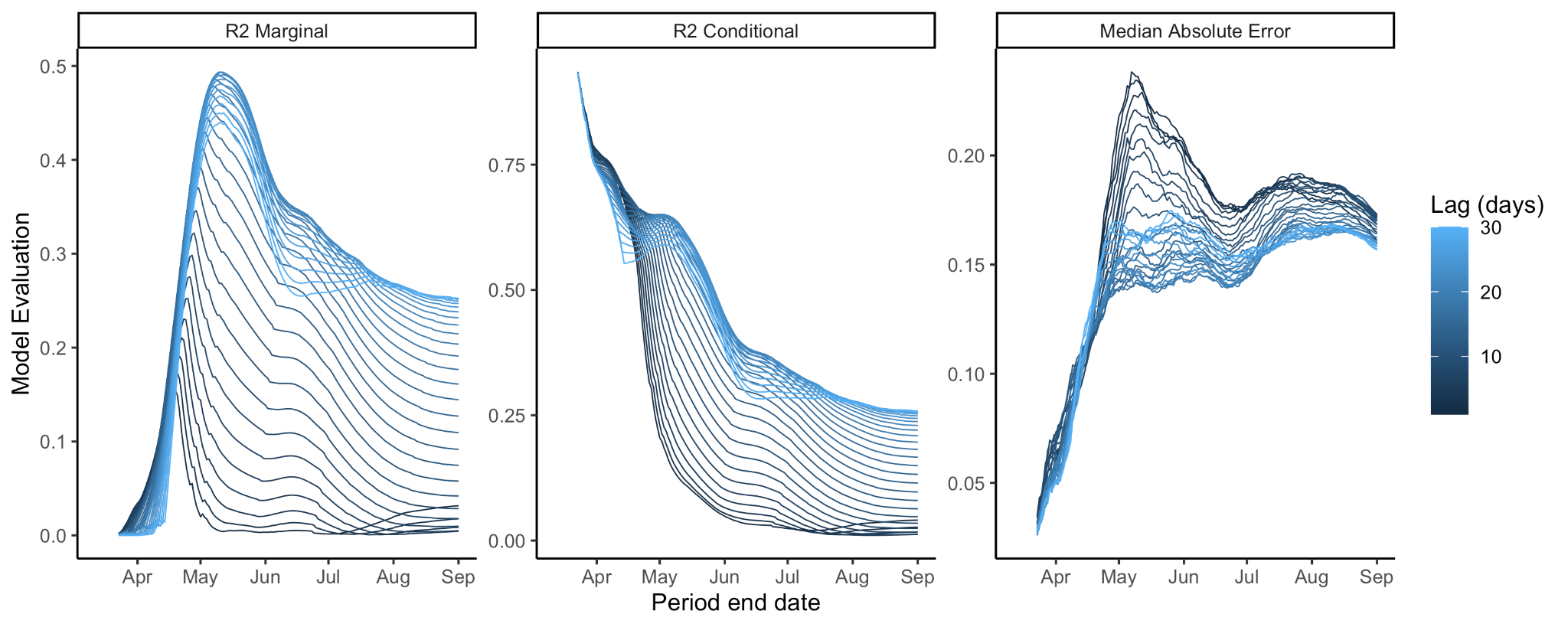

**Supplemental Figure 12. *Model performance for different periods.*** *The performance of the multilevel model through time, measured by marginal and conditional R^2^ (trained on periods beginning 12th March and ending from 19th March to 1st September).*

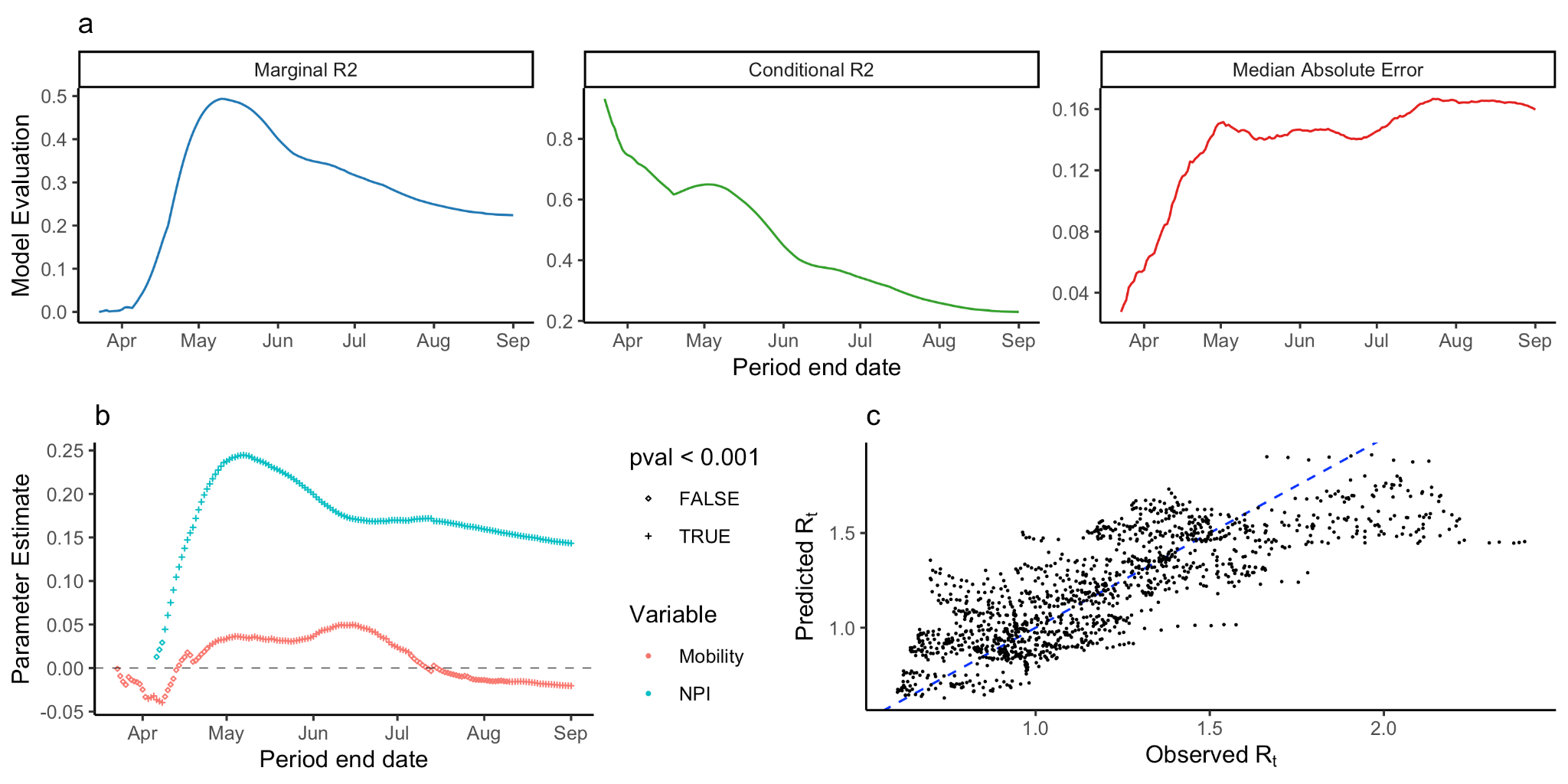

**Supplemental Figure 13. *Statistical analysis of R_t_.*** *a) The marginal R^2^ of the multilevel model trained on varying-length periods through time. b) Observed vs Predicted R_t_ for the model trained between 12th March and 11th May, 2020. Blue dashed line shows diagonal where x = y. c) The maximum marginal R^2^ for all periods trained for different lag values from 0 to 30 days.*

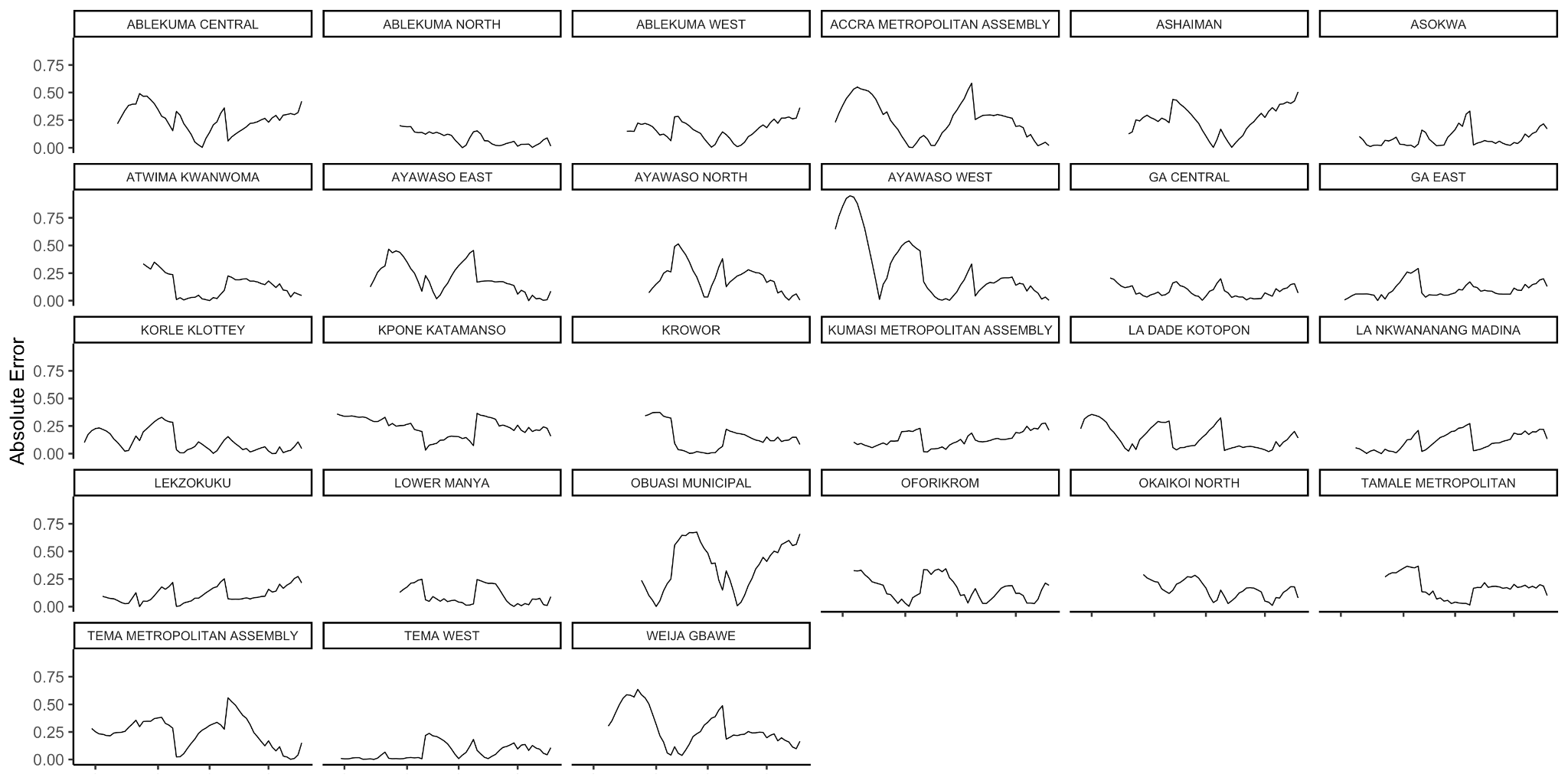

***Supplemental Figure 14. Absolute error by date and district.*** *The absolute error of model estimations on each date per district.*

| **Predictors** | **Estimates** | **CI** | **p** |
| --- | --- | --- | --- |
| (Intercept) | 1.196 | 1.142 – 1.250 | <0.001 |
| NPI | 0.242 | 0.228 – 0.256 | <0.001 |
| Mobility Residuals | 0.035 | 0.020 – 0.050 | <0.001 |
| Holidays | -0.020 | -0.084 – 0.044 | 0.540 |

**Supplemental Table 3. *Regression coefficients for the multilevel model.*** *Regression coefficients for the multilevel model trained between 12th March and 11th May, 2020. Table shows coefficients, 95% confidence intervals, and p values for each predictor.*

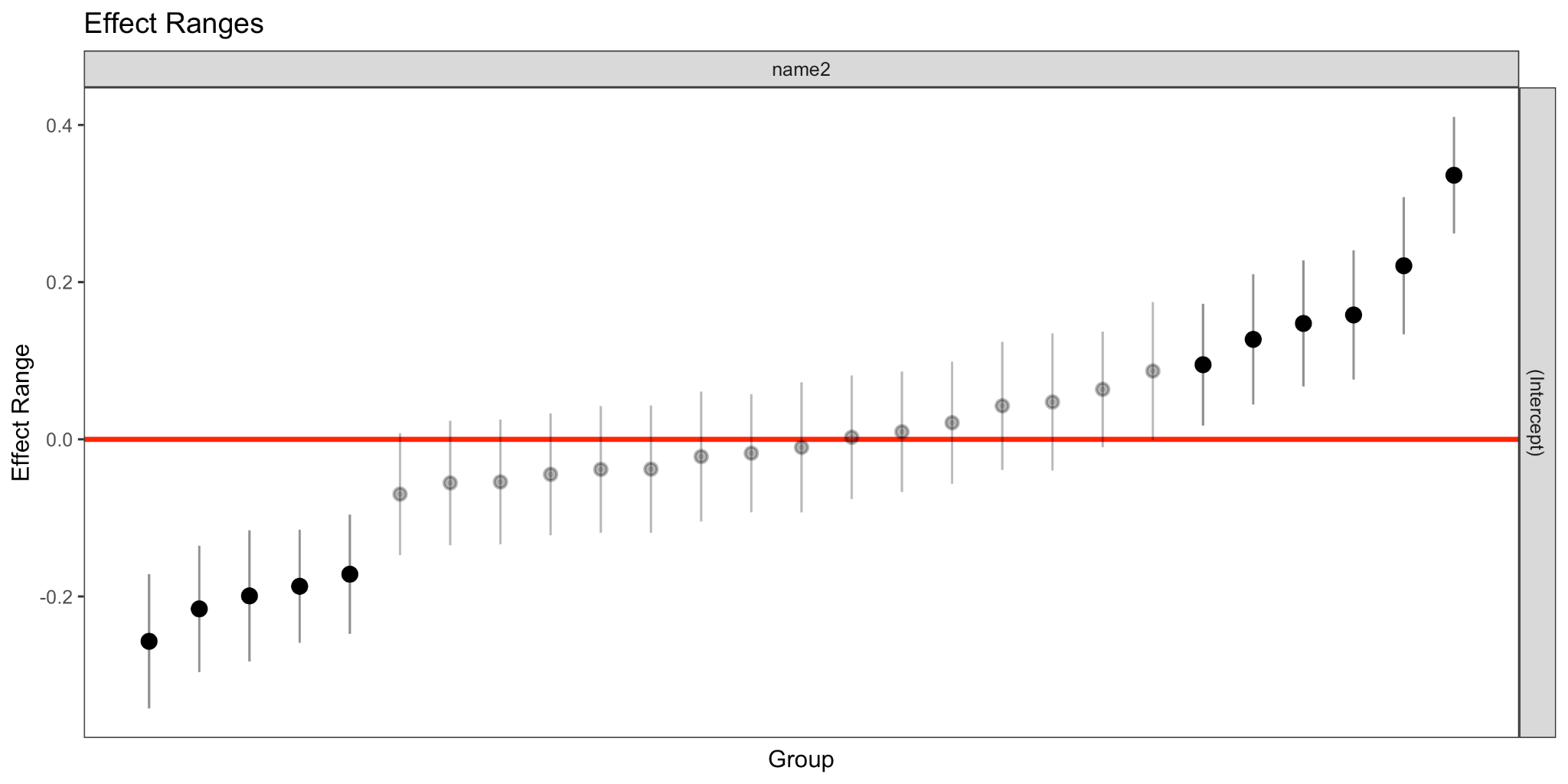

**Supplemental Figure 15. *Random effect sizes for a model with truncated time series.*** *The district-specific random effect sizes for the model trained between 12th March and 11th May. Eleven random effects are distinguishable from 0.*

**Supplemental Section 3**

Sensitivity analysis of model outputs using the Google Mobility Indicator and custom stringency index. This analysis shows similar results to the main with an increase in the model’s explanation of R_t_ variance early in the epidemic and subsequent decrease after June 2020 (Supplemental Figure 14). Google mobility has limited spatial extent and does not include all the districts included in the main analysis. In this analysis we identified an optimal lag of 22 days (Supplemental Figure 19).

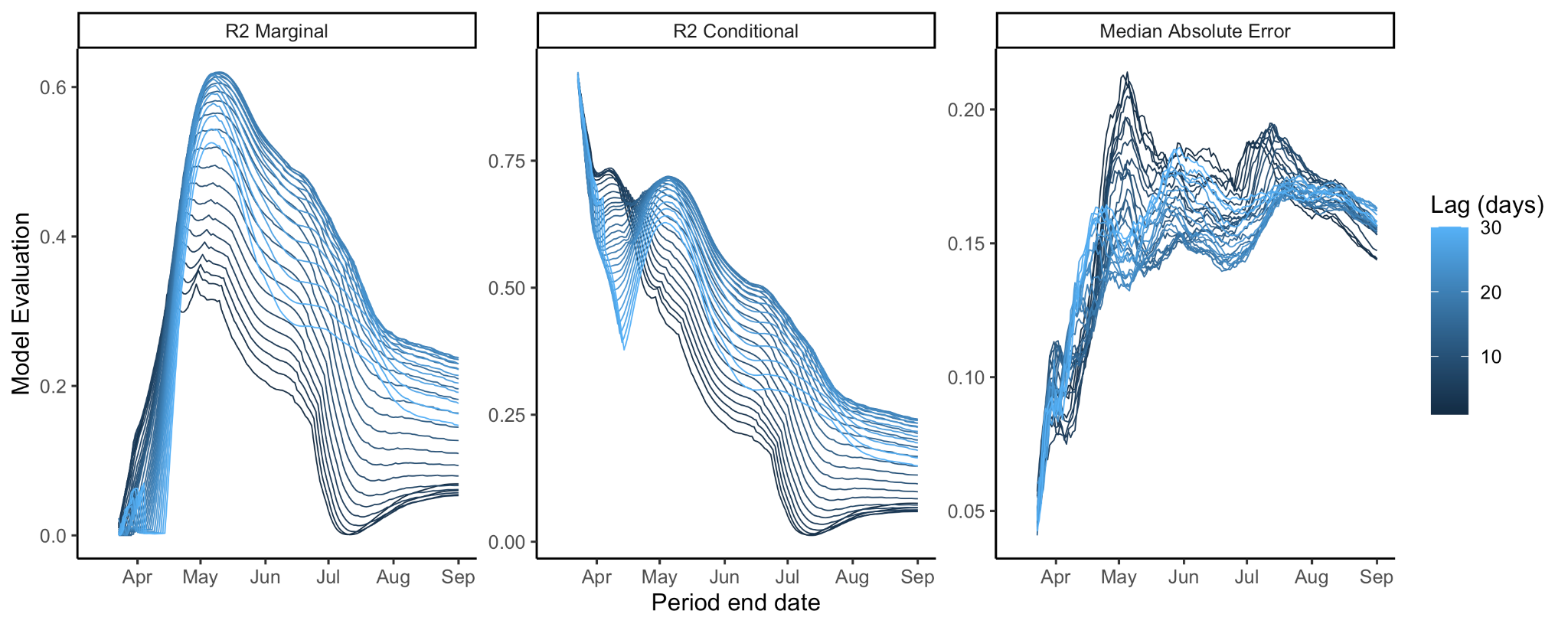

**Supplemental Figure 16. *Model performance for different periods.*** *The performance of the multilevel model through time, measured by marginal and conditional R^2^ (trained on periods beginning 12th March and ending from 19th March to 1st September).*

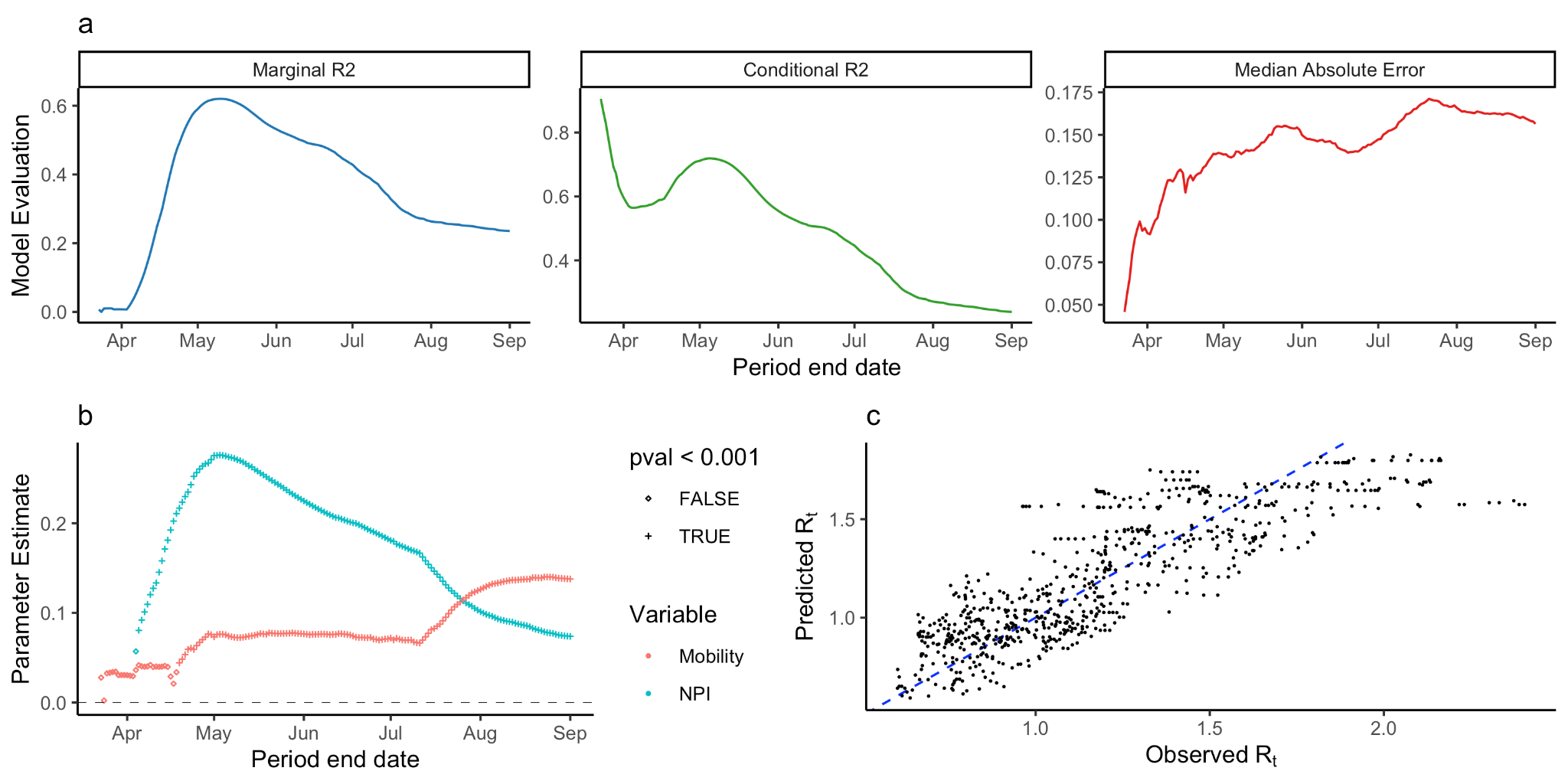

**Supplemental Figure 17. *Statistical analysis of R_t_.*** *a) The marginal R^2^ of the multilevel model trained on varying-length periods through time. b) Observed vs Predicted R_t_ for the model trained between 12th March and 10th May, 2020. Blue dashed line shows diagonal where x = y. c) The maximum marginal R^2^ for all periods trained for different lag values from 0 to 30 days.*

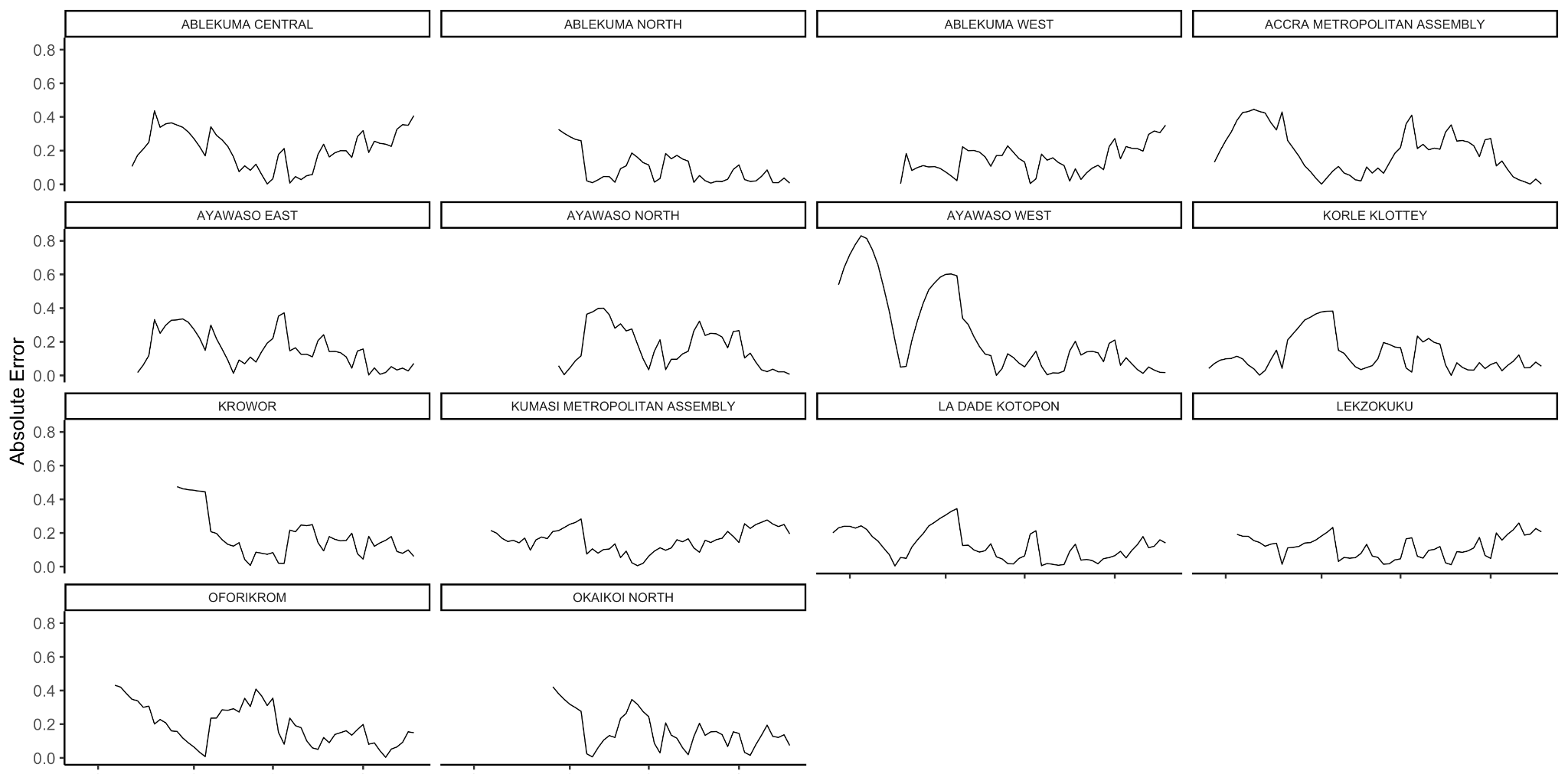

***Supplemental Figure 18. Absolute error by date and district.*** *The absolute error of model estimations on each date per district.*

| **Predictors** | **Estimates** | **CI** | **p** |
| --- | --- | --- | --- |
| (Intercept) | 1.201 | 1.136 – 1.265 | <0.001 |
| NPI | 0.270 | 0.252 – 0.288 | <0.001 |
| Mobility Residuals | 0.072 | 0.054 – 0.091 | <0.001 |
| Holidays | 0.008 | -0.078 – 0.093 | 0.861 |

**Supplemental Table 4. *Regression coefficients for the multilevel model.*** *Regression coefficients for the multilevel model trained between 12th March and 10th May, 2020. Table shows coefficients, 95% confidence intervals, and p values for each predictor.*

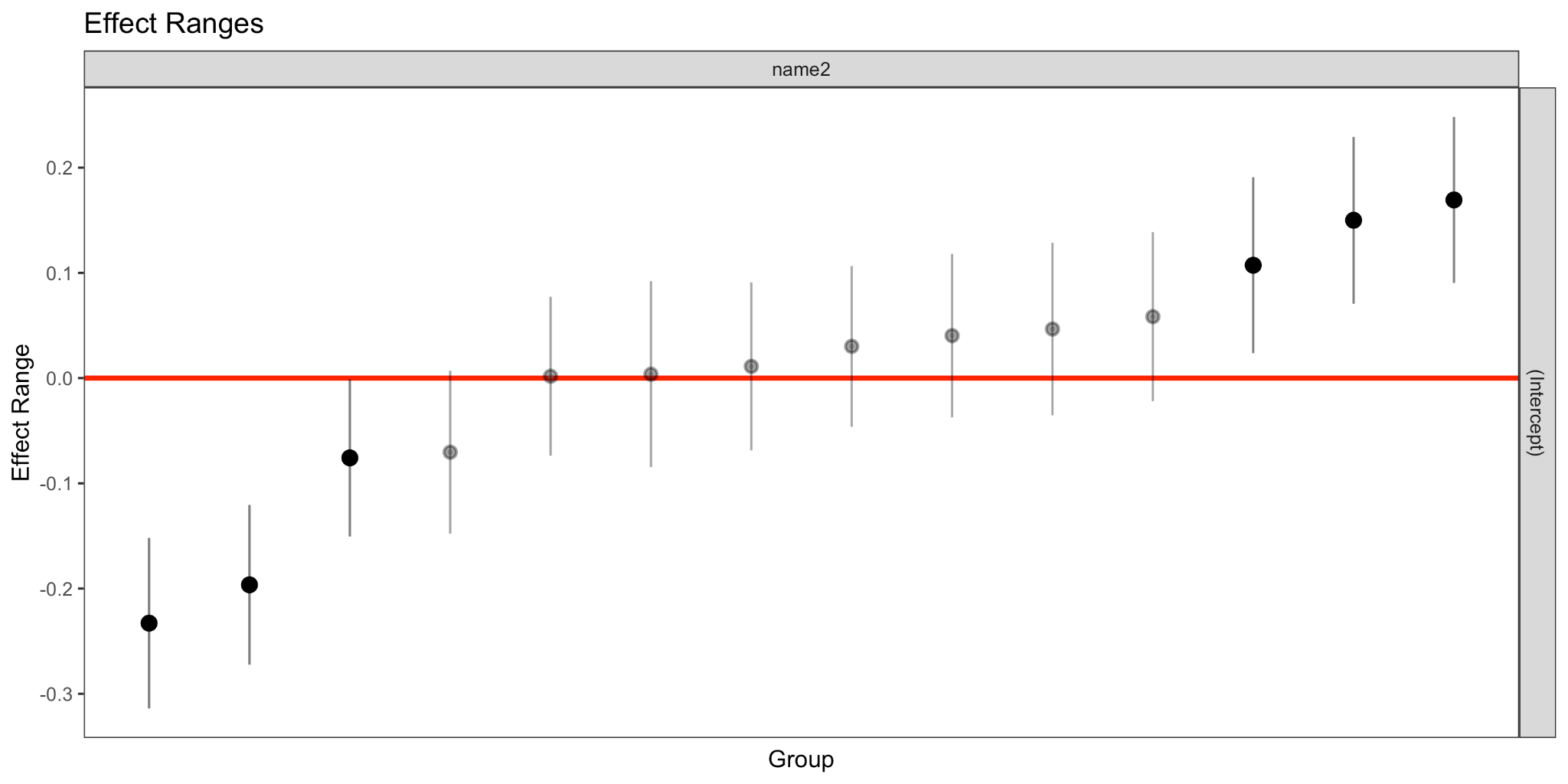

**Supplemental Figure 19. *Random effect sizes for a model with truncated time series.*** *The district-specific random effect sizes for the model trained between 12th March and 10th May. Five random effects are distinguishable from 0.*

**Supplemental Section 4**

Sensitivity Analysis with fixed time windows (30, 60, 90 days). Instead of extending the time period of the model (as in the main text) we used a fixed-length sliding window to assess changes in fit at different times of the epidemic. We found a similar pattern of model performance early in the epidemic as observed in the main model. Later in the epidemic, we found that the performance of the model was sensitive to the length of the time period included. With the 60 day period, for example, the model performance increased during July. Because this increase is diminished when using longer rolling periods, this likely reflects the alignment of the 60 day window with a period of reduced variance in R_t_.

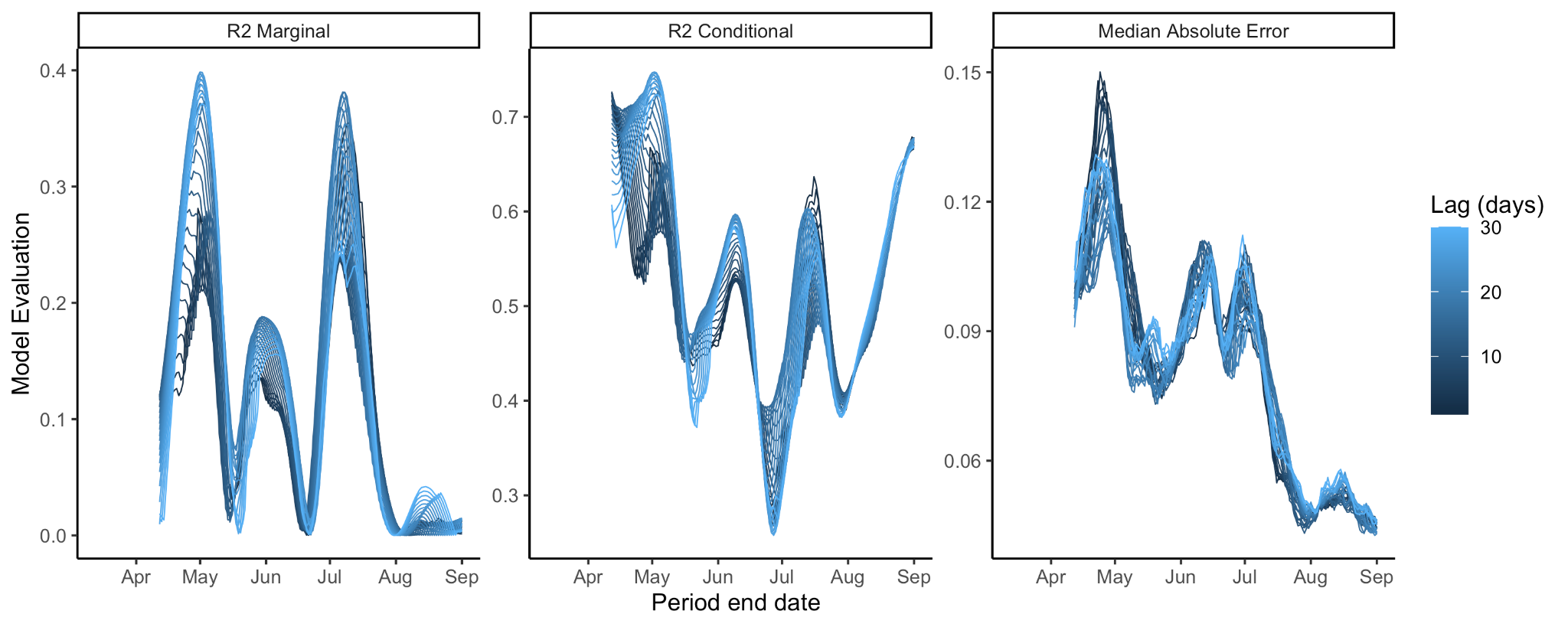

**Supplemental Figure 20. *Model performance for different periods with a 30 day sliding time window*** *The performance of the multilevel model through time, measured by marginal and conditional R^2^ (trained on rolling 30 day periods between 12th March and 1st September).*

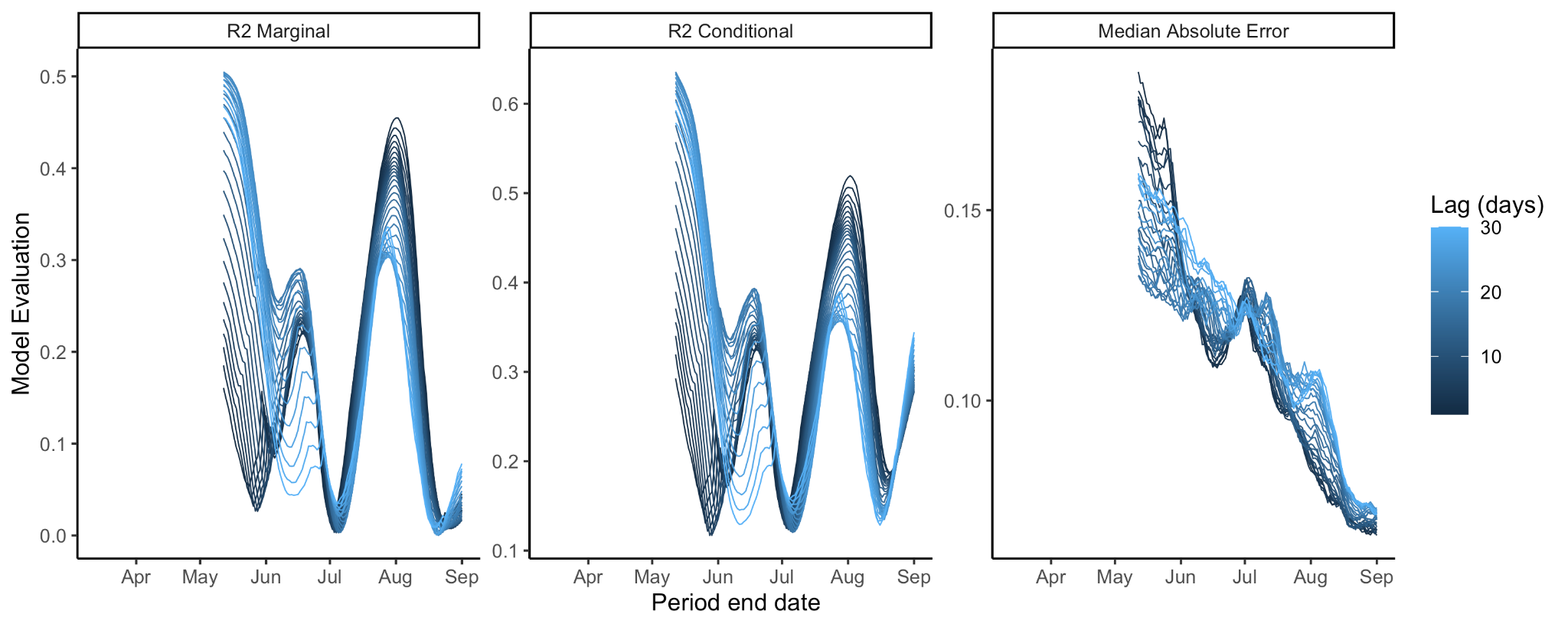

**Supplemental Figure 21. *Model performance for different periods with a 60 day sliding time window.*** *The performance of the multilevel model through time, measured by marginal and conditional R^2^ (trained on rolling 60 day periods between 12th March and 1st September).*

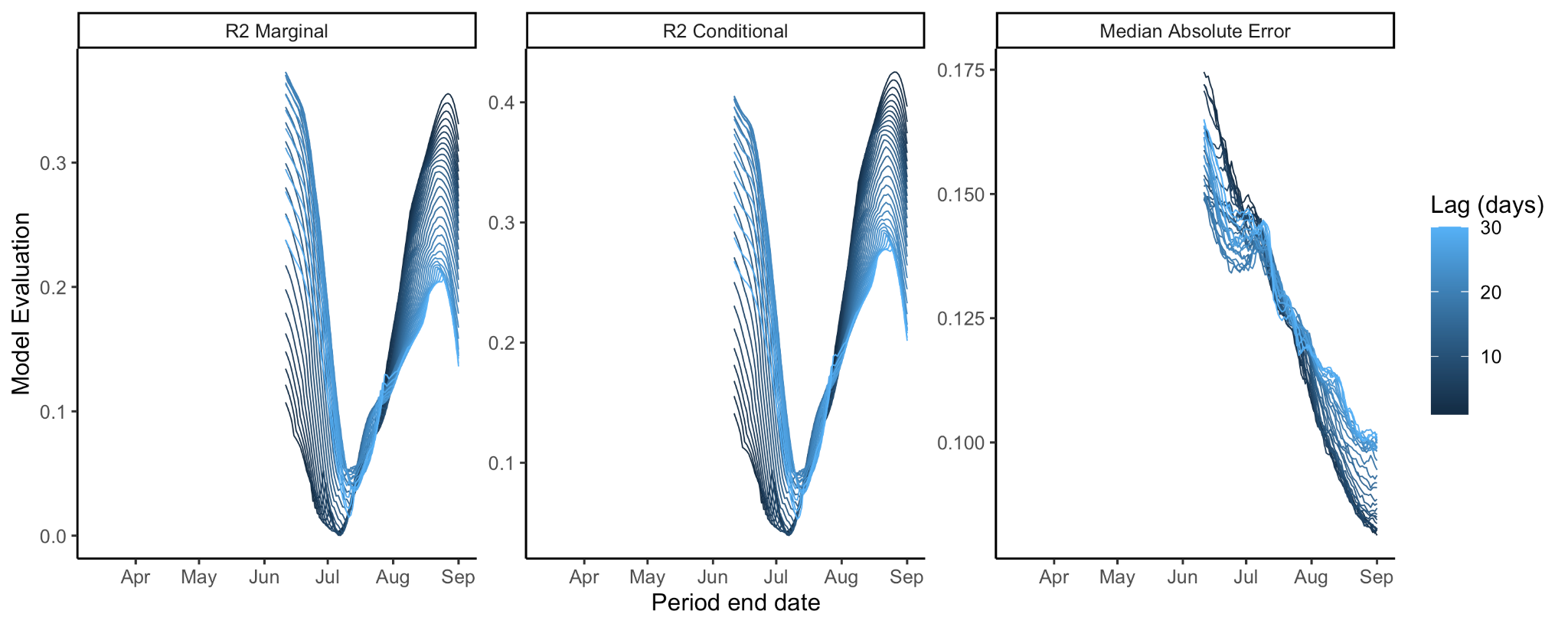

**Supplemental Figure 22. *Model performance for different periods with a 90 day sliding time window.*** *The performance of the multilevel model through time, measured by marginal and conditional R^2^ (trained on rolling 90 day periods between 12th March and 1st September).*
